# Effects of sex, treatment resistance, and antidepressant treatment on monoamine oxidase B total distribution volume

**DOI:** 10.64898/2026.09.08.26362462

**Authors:** Emily JY Kuik, Catalina Baena-Tan, Sho Moriguchi, Joeffre Braga, Laura Miler, Pablo Rusjan, Michael Bagby, M Ishrat Husain, Neil Vasdev, Toshifumi Tomoda, Mounira Banasr, Stefan Kloiber, Jeffrey H Meyer

## Abstract

Monoamine oxidase B (MAO-B) is a high-density protein located mainly in astrocytes that influences mitochondrial function and transition to astrogliosis. MAO-B produces hydrogen peroxide while metabolizing non-serotonergic monoamines. Elevated MAO-B level is implicated in the pathophysiology of several common neuropsychiatric diseases. The aims are to examine the effect of treatment resistant major depressive episodes, sex, and antidepressant treatment on MAO-B total distribution volume ([^11^C]SL25.1188 V_T_), index of MAO-B level. [^11^C]SL25.1188 PET was applied in 95 adults (29 with MDE and no history of treatment-resistance (NTR-MDE), 26 with MDE and history of treatment-resistance (TRD), and 40 healthy control (HC)). A subset of NTR-MDE and TRD (n=13) were scanned before and after treatment with phenelzine, rasagiline, or duloxetine. [^11^C]SL25.1188 V_T_ was higher in grey matter regions in females (10%, *p*<.001). [^11^C]SL25.1188 V_T_ was also higher in prefrontal cortex in TRD compared to HC (23%, p<.001) and TRD compared to NTR-MDE (6%, *p*=.033). Duloxetine had minimal effect on [^11^C]SL25.1188 V_T_, but occupancy of rasagiline and phenelzine was ∼94%. This study discovered sex differences in MAO-B level, which may explain sex differences in prevalence or trajectory of illnesses with greater MAO-B level like Alzheimer’s disease, traumatic brain injury and major depressive disorder. Given our findings of greater MAO-B level in prefrontal cortex of TRD and its insensitivity to serotonin reuptake inhibition, combined with potential harm of highly elevated MAO-B level, development of MAO-B inhibitors could be considered for a subset of TRD. Rasagiline and phenelzine demonstrate similarly potent occupancy in vivo at clinical dosage.

## Introduction

Monoamine oxidase B (MAO-B), a high-density enzyme in the human brain (1.8 to 34 pmol/mg in grey matter), is primarily located on the outer mitochondrial membrane in astrocytes as well as the cell bodies of serotonergic and dopaminergic neurons [1, 2]. In brain tissue, MAO-B level is of considerable interest because it is highly correlated with MAO-B activity [2, 3]. MAO-B is an ongoing target of drug development [4, 5], as it plays important roles in the pathophysiology of psychiatric and neurological diseases. Illnesses with greater MAO-B level include the prefrontal cortex (PFC) of major depressive disorder (MDD) [6], the grey matter of Alzheimer’s disease [7], the grey matter of long COVID [8], the cortex of traumatic brain injury with chronic persistent symptoms [9], and the midbrain of Parkinson’s disease [10]. Previous studies demonstrated a number of mechanisms by which MAO-B mediates pathophysiological consequences. MAO-B produces reactive oxygen species (i.e., hydrogen peroxide) while metabolizing non-serotonergic monoamines, such as dopamine, norepinephrine, and phenylethylamine [11, 12]. Greater production of reactive oxygen species could account for negative impacts of overexpressed MAO-B on mitochondrial complex I activity [12] and hence oxidative phosphorylation. MAO-B also contributes to the development and persistence of astrogliosis, a pathologically activated state of glial fibrillary acidic protein (GFAP)-positive astrocytes [12–15]. In addition, stress and the stress-associated steroid hormones, glucocorticoids, upregulate MAO-B transcription via direct binding of the glucocorticoid receptor to the glucocorticoid response element 4 (GRE4) in the MAO-B promoter. An alternative mechanism of glucocorticoid effect involves transcriptional repressors (i.e., E2F-associated phosphoprotein (EAPP) and R1), which compete with a transcriptional activator, Simian virus promoter factor 1 (Sp1), for binding to the Sp1 sites in the MAO-B promoter region. Under stress conditions, reduced occupancy of EAPP and R1 at the Sp1 site in the MAO-B promoter therefore leads to elevated level of MAO-B and hence greater MAO-B activity [16–18].

[^11^C]SL25.1188 V_T_, an index of MAO-B level, may be measured *in vivo* in humans applying the positron emission tomography (PET) radiopharmaceutical [^11^C]SL25.1188. This radiopharmaceutical has outstanding qualities, including very high specific binding to non-displaceable binding ratio (>8), excellent reversibility, high brain uptake, selectivity for MAO-B, and an absence of brain-penetrant radioactive metabolites [19, 20].

The first aim of this study is to investigate whether there are differences in PFC MAO-B V_T_ during major depressive episodes with a history of treatment resistance (TRD) compared to major depressive episodes with no history of treatment resistance (NTR-MDE). Greater PFC MAO-B V_T_ was previously found during MDE of MDD, and MAO-B is not targeted directly by first- and second-line serotonin and serotonin and norepinephrine reuptake inhibitor medication treatments [21]. It is hypothesized that the highest level of PFC MAO-B V_T_ is associated with TRD because overexpression of MAO-B is associated with greater removal of non-serotoninergic monoamines, greater production of reactive oxygen species, and loss of mitochondrial complex I [11, 12]. Each of these sequelae are also not direct targets of serotonin reuptake inhibitor treatments, yet may contribute to MDE symptoms: In humans, depletion of non-serotonergic monoamines, such as through α-methylparatyrosine administration, may rapidly induce depressive symptoms [22]. Greater production of reactive oxygen species has been proposed to create risk for depressive symptoms [23, 24] and genetic disorders with loss of mitochondrial complex I level or activity are associated with high risk of depressive symptoms [25, 26].

The second aim of this study is to determine whether there are any differences in MAO-B V_T_ in females compared to males across grey matter regions, including the PFC, anterior cingulate cortex (ACC), dorsal putamen, ventral striatum, thalamus, and hippocampus. This study measures an index of MAO-B level, and MAO-B level is consistently correlated with MAO-B activity in brain tissue, yet the effect of sex on either had not been prioritized in previous studies of the human brain. The present study is better suited to address this question due to its sampling, being balanced between males and females, and of relatively large sample size, including about twice as many females as the largest samples among previous studies. Since estrogens lower the expression of MAO-B transcription by downregulating the expression of oestrogen-related receptors, which constitutively upregulate MAO-B gene transcription [27, 28], it is hypothesized that MAO-B V_T_ will be lower in females in the grey matter regions prioritized. In addition, as mechanisms to account for potential differences in MAO-B V_T_ between sex have had minimal investigation, we also explored gene expression differences between females and males in a publicly-available microarray dataset derived from postmortem human brain tissue, focusing on those related to MAO-B expression.

The third aim of this study is to investigate the effects of rasagiline, an MAO-B inhibitor; phenelzine, an MAO-A and MAO-B inhibitor; and duloxetine, a selective serotonin and norepinephrine reuptake inhibitor (SNRI), on MAO-B V_T_, and characterize MAO-B occupancy for the first two medications. Occupancy may be derived from V_T_ measurements in which there is a substantial lowering in V_T_ at the time of the second PET scan, which is expected for the first two medications [29]. Rasagiline occupancy is of interest to justify potential repurposing because it is a well-tolerated, selective, and irreversible MAO-B inhibitor that typically does not require the dietary restriction of low tyramine intake for inhibitors of both MAO-A and MAO-B [30].

Tyramine is metabolized by both MAO-A and MAO-B, so substantial inhibition of only one enzyme still allows adequate metabolism of tyramine, whereas medications with substantial inhibition of both MAO-A and MAO-B require dietary restriction to avoid hypertensive crisis from elevated plasma tyramine level [11]. Rasagiline is primarily used for Parkinson’s disease and has not been advanced for repurposing in other illnesses [30]. Phenelzine represents a positive control, as it is a nonselective irreversible MAO inhibitor that has an indication for MDE of MDD but is not commonly used because it requires dietary restriction of tyramine and cannot be given with other monoamine-influencing antidepressants [31]. Other advantages of determining phenelzine occupancy as compared to the other option of tranylcypromine is that the occupancy of phenelzine for MAO-A is known at common treatment dose, and phenelzine is sufficiently well-tolerated to reach the clinical dosing associated with its initial development in open trial, whereas tranylcypromine is difficult to tolerate at dosage supported by clinical trial evidence [32–34]. Non-quantitative assessment of rasagiline with [^11^C]deprenyl PET suggests that rasagiline has detectable occupancy [35], but the present study will provide quantitative results. Duloxetine is anticipated to be a negative control as it is classified as a SNRI with no direct MAO-B inhibition or binding.

## Patients and methods

### Participants

Participants were either healthy, had NTR-MDE or had TRD. For healthy participants, the main inclusion criteria was no history of psychiatric illness based on the Structured Clinical Interview for DSM-4 or DSM-5 (SCID) [36] and good physical health based on a structured health questionnaire. The key inclusion criteria for NTR-MDE and TRD participants included primary diagnosis of MDE with early-onset MDD, with first MDE prior to age 40, verified by the SCID for DSM-4 or DSM-5 [36] and a score of greater than or equal to 17 on the 17-item Hamilton Depression Rating Scale (HDRS). Categorization as TRD versus NTR-MDE was additionally based on history of non-response to at least two different classes of antidepressants or at least one SNRI. Important exclusion criteria included use of antidepressants within the past 4 weeks for NTR-MDE and TRD, and lifetime use for healthy. Exclusion criteria common to all participants were history of psychotic symptoms, antisocial or borderline personality disorders based on the Structured Clinical Interview for DSM-IV (SCID-II) [37] or Structured Clinical Interview for DSM-5 Personality Disorders Version (SCID-5-PD), and neurodegenerative illness; cigarette smoking for the past 6 months or current use of recreational drugs with recent use verified by urine drug screen; current alcohol use disorder; electroconvulsive therapy or mechanical brain stimulation treatment within the previous 6 months; and use of MAO-B inhibitor treatments within the last 4 weeks.

Additional inclusion and exclusion criteria can be found in Supplementary Materials. Participants provided written informed consent after all procedures were fully explained, and all components of the study were approved by the Research Ethics Board of the Centre for Addiction and Mental Health (CAMH) and Health Canada.

### [^11^C]25.1188 V_T_ Measurement

PET imaging data was acquired from each participant for 90 minutes with a 3D high-resolution research tomograph (HRRT; CPS/Siemens) PET scanner system as previously described [20]. Arterial sampling was done concurrently during the PET scan using an automatic blood sampling system and manual sampling during the emission PET scan. An MRI image (Discovery MR750 3T GE scanner) was acquired for delineation of regions of interest (ROI). [^11^C]SL25.1188 V_T_ was calculated using a 2-tissue compartment model previously validated for [^11^C]SL25.1188 PET [20]. Both PET and MRI scans were completed in Toronto. See Supplementary Materials for additional details.

### Treatment Interventions on [^11^C]SL25.1188 V_T_

A subset of participants also partook in the open trial treatment portion of the study. NTR-MDE received duloxetine. Assignment of TRD participants was substantially consistent with usual treatment algorithms, such that participants received duloxetine if they had no history of non-response to SNRI duloxetine or venlafaxine. Otherwise, TRD participants received phenelzine if they felt they could follow the dietary restriction requirement for phenelzine or rasagiline if they felt they could not follow the dietary restriction requirement for phenelzine. Administration of medication was open label and began the day following the first [^11^C]SL25.1188 PET scan.

Phenelzine was administered at 15 mg once daily and raised every 4 to 7 days by 15 mg to achieve a total daily dose of 45 to 60 mg given over two doses based on tolerability. Rasagiline was administered at 0.5 mg daily for 4 to 7 days, then raised to 1 mg daily for 6 to 8 weeks if well tolerated. Otherwise, dosing remained at 0.5 mg daily. Duloxetine was given at 30 mg daily in week 1, then raised to 60 mg daily for 6 weeks. The follow-up PET scan occurred 4 to 6 weeks after titration to the optimized dose and was further timed to avoid peak plasma level for the MAO-B inhibitors (i.e., scanning occurred 6 to 24 hours after last dose for each drug). Non-responders to medication received a tapering dose for one week before the medication was stopped (unless they were already taking the minimum dose of rasagiline at 0.5 mg daily). Responders were given the option to continue treatment at the dose associated with response, with a transition to their regular treating physician. Compliance was assessed via pill count done halfway through the trial and at the end of the trial.

### Exploration of Sex Differences in mRNA

Microarray data for 18 females and 20 males from a sample of individuals with major depressive disorder (n=19) and controls (n=19) from the Gene Expression Omnibus dataset GSE53987, sampling the hippocampus, dorsolateral prefrontal cortex, and associative striatum regions [38, 39], was used to explore the presence of sex differences in the expression of several mRNA related to the MAO-B protein level, including MAO-B, regulators of MAO-B transcription (EAPP, R1), astrogliosis (GFAP), and mitochondrial density (citrate synthase) (Supplemental Materials).

### Statistical Analyses

The analysis determining the effect of sex on [^11^C]SL25.1188 V_T_ was a linear mixed-effects model (LMM) evaluating sex, group, and region (PFC, ACC, dorsal putamen, ventral striatum, thalamus, hippocampus) as fixed effects and participant as a random effect, with [^11^C]SL25.1188 V_T_ as the dependent variable. Sex was assessed first so as to consider its inclusion in the comparisons of illness groups. The analysis comparing [^11^C]SL25.1188 V_T_ in PFC between TRD and NTR-MDE applied two similar methodologically overlapping approaches. One was an ANCOVA with group and sex as predictor variables and whole PFC [^11^C]SL25.1188 V_T_ as the dependent variable. The second analysis applied a LMM evaluating sex, group, region (dorsolateral PFC, ventrolateral PFC, medial PFC, and orbitofrontal cortex), and group*region as fixed effects and participant as a random effect, with [^11^C]SL25.1188 V_T_ as the dependent variable.

To assess whether there was a change in [^11^C]SL25.1188 V_T_ before and after antidepressant treatment, the impact for each drug was determined separately by applying a LMM evaluating time and region (PFC, ACC, dorsal putamen, ventral striatum, thalamus, hippocampus) as fixed effects and participant as a random effect, with regional [^11^C]SL25.1188 V_T_ as the repeated dependent variable. For interventions with a substantial change of greater than 30% in [^11^C]SL25.1188 V_T_, a Lassen plot was applied to determine a mean occupancy of [^11^C]SL25.1188 V_T_ across brain regions and a 95% confidence interval for occupancy was found for each intervention for which a substantive change in [^11^C]SL25.1188 V_T_ was present.

## Results

### Participants

Ninety-five participants, age 18 to 65 years (52 females, 43 males), were included in the analyses. Of these, 40 were healthy, 29 had NTR-MDE and 26 had TRD. Age and sex were not significantly different across all three groups (Table 1). There were no significant differences in HDRS score at screening, number of previous MDEs, and duration of illness across NTR-MDE and TRD groups (Table 1).

**Table 1.** Demographic and Clinical Variables.

|  | Participants |  |  | *p* |
| --- | --- | --- | --- | --- |
|  | NTR-MDE<br>(*n* = 29) | TRD (*n* = 26) | Healthy<br>Controls<br>(*n* = 40) |  |
| Age (years) | 31.3 ± 10.4 | 35.7 ± 12.9 | 31.9 ± 10.9 | .294 |
| Sex (Female:Male) | 18:11 | 11:15 | 23:17 | .305 |
| Hamilton Depression<br>Rating Scale | 19.0 ± 4.2 | 19.9 ± 3.7 | - | .428 |
| Number of past<br>MDEs<sup>i</sup> | 4.3 ± 3.0 | 6.1 ± 3.4 | - | .055 |
| Age of onset (years) | 23.5 ± 8.6 | 20.9 ± 8.7 | - | .323 |

Data presented as mean ± standard deviation, unless stated otherwise.
Statistical analyses were performed using analysis of variance (ANOVA), with the exception of sex, which was performed using $\chi^2$ test.
<sup>i</sup>The maximum number of MDEs counted is 10.

### Effect of sex on [^11^C]SL25.1188 V_T_

[^11^C]SL25.1188 V_T_ was significantly higher in females compared to males in the PFC, ACC, ventral striatum, dorsal putamen, thalamus and hippocampus (LMM; *F*_1,86_ = 17.57*, p* <.001; Fig. 1 and Table 2). An additional analysis extending to include more grey matter regions (parietal cortex, occipital cortex, temporal cortex and midbrain) with the same predictor variables also found that [^11^C]SL25.1188 V_T_ was significantly elevated in females (LMM; *F*_1,88_ = 16.30*, p* <.001; Table 2).

**Figure 1:**
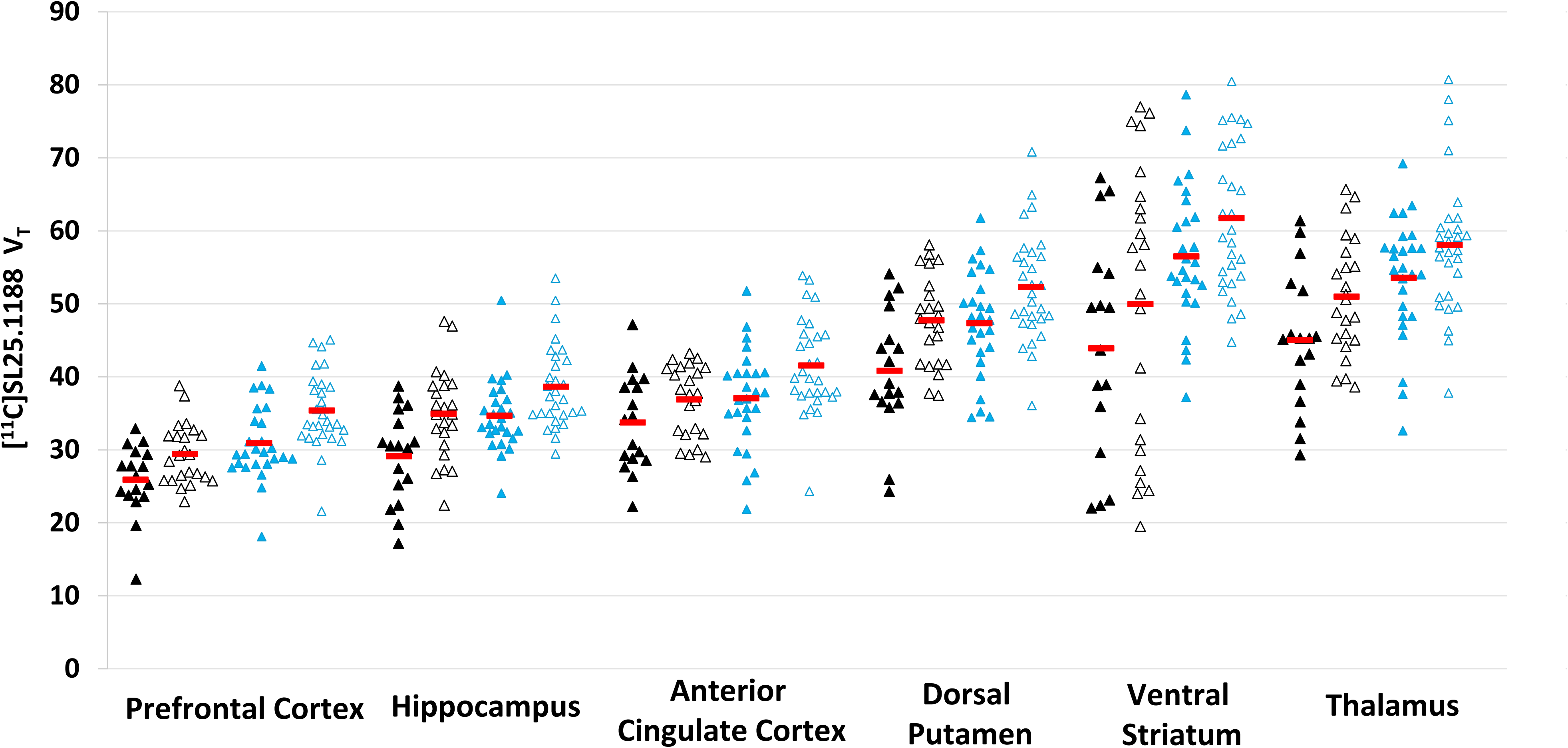
[^11^C]SL25.1188 total distribution volume (V_T_) in brain regions of interest in females vs. males. A linear mixed-effects model found an effect of sex on [^11^C]SL25.1188 V_T_, with greater [^11^C]SL25.1188 V_T_ in females (*F*_1,86_ = 17.57*, p* <.001). Filled triangles represent males (*n* = 43). Hollow triangles represent females (*n* = 52). Black triangles represent healthy controls (*n* = 40). Blue triangles represent MDE participants, which includes both NTR-MDE and TRD participants (*n* = 29). Red horizontal bars represent means.

**Table 2.** Regional [^11^C]SL25.1188 V_T_ in females and males.

| Region of Interest | Females<br>(*n* = 52) |  | Males<br>(*n* = 43) |  | Difference | *F* | *p* |
| --- | --- | --- | --- | --- | --- | --- | --- |
|  | Mean | SD | Mean | SD |  |  |  |
| Prefrontal Cortex | 32.7 | 5.6 | 28.9 | 5.5 | 12.3% | 11.352 | .001 |
| Dorsolateral Prefrontal Cortex | 32.7 | 5.2 | 28.6 | 6.2 | 13.4% | 12.517 | <.001 |
| Ventrolateral Prefrontal Cortex | 29.7 | 5.1 | 26.4 | 5.4 | 11.8% | 9.477 | .003 |
| Medial Prefrontal Cortex | 36.4 | 6.0 | 32.7 | 6.1 | 10.7% | 8.668 | .004 |
| Orbitofrontal Cortex | 29.6 | 5.7 | 27.3 | 4.9 | 8.1% | 4.410 | .038 |
| Anterior Cingulate Cortex | 39.5 | 6.2 | 35.7 | 6.8 | 10.1% | 7.944 | .006 |
| Hippocampus | 37.0 | 6.2 | 32.5 | 6.1 | 13.0% | 13.023 | <.001 |
| Dorsal Putamen | 50.3 | 7.2 | 44.7 | 8.3 | 11.8% | 12.339 | <.001 |
| Ventral Striatum | 60.3 | 12.7 | 53.6 | 11.2 | 11.8% | 7.290 | .008 |
| Thalamus | 54.9 | 9.5 | 50.2 | 9.6 | 8.9% | 5.824 | .018 |
| Midbrain | 37.9 | 7.1 | 34.6 | 6.7 | 9.1% | 5.404 | .022 |
| Parietal Cortex | 31.8 | 4.7 | 29.3 | 5.4 | 8.2% | 5.933 | .017 |
| Temporal Cortex | 31.9 | 4.8 | 29.8 | 6.0 | 6.8% | 3.677 | .058 |
| Occipital Cortex | 27.3 | 3.9 | 25.0 | 4.4 | 8.8% | 7.337 | .008 |

Means and standard deviations presented in table are derived from raw unadjusted $V_T$ values. P values are derived from an analysis of variance (ANOVA).

### Effect of TRD versus NTR-MDE or Health on [^11^C]SL25.1188 V_T_

[^11^C]SL25.1188 V_T_ in PFC was significantly elevated in TRD compared to NTR-MDE (ANCOVA, factors of group and sex; effect of group: *F*_1,52_ = 4.78*, p* = 0.033; Fig. 2 and Table S1). Similar results were found comparing [^11^C]SL25.1188 V_T_ in TRD to NTR-MDE across subregions of the PFC, as there was a group effect but no subregion-specific difference (LMM; predictors of group, region, sex, group by region; effect of group: *F*_1,52_ = 4.45, *p* = .040; Fig. 2 and Table S1). Consistent with these findings, [^11^C]SL25.1188 V_T_ in PFC was significantly elevated in TRD compared to the healthy group (ANCOVA, factors of group and sex; effect of group: *F*_1,63_ = 35.39, *p* < 0.001; Fig. 2 and Table S2). There was also an effect of group comparing subregions of the PFC but no subregion-specific findings (LMM; predictors of group, region, sex, group by region; effect of group: *F*_1,63_ = 19.23*, p* < 0.001; Fig. 2 and Table S2).

**Figure 2:**
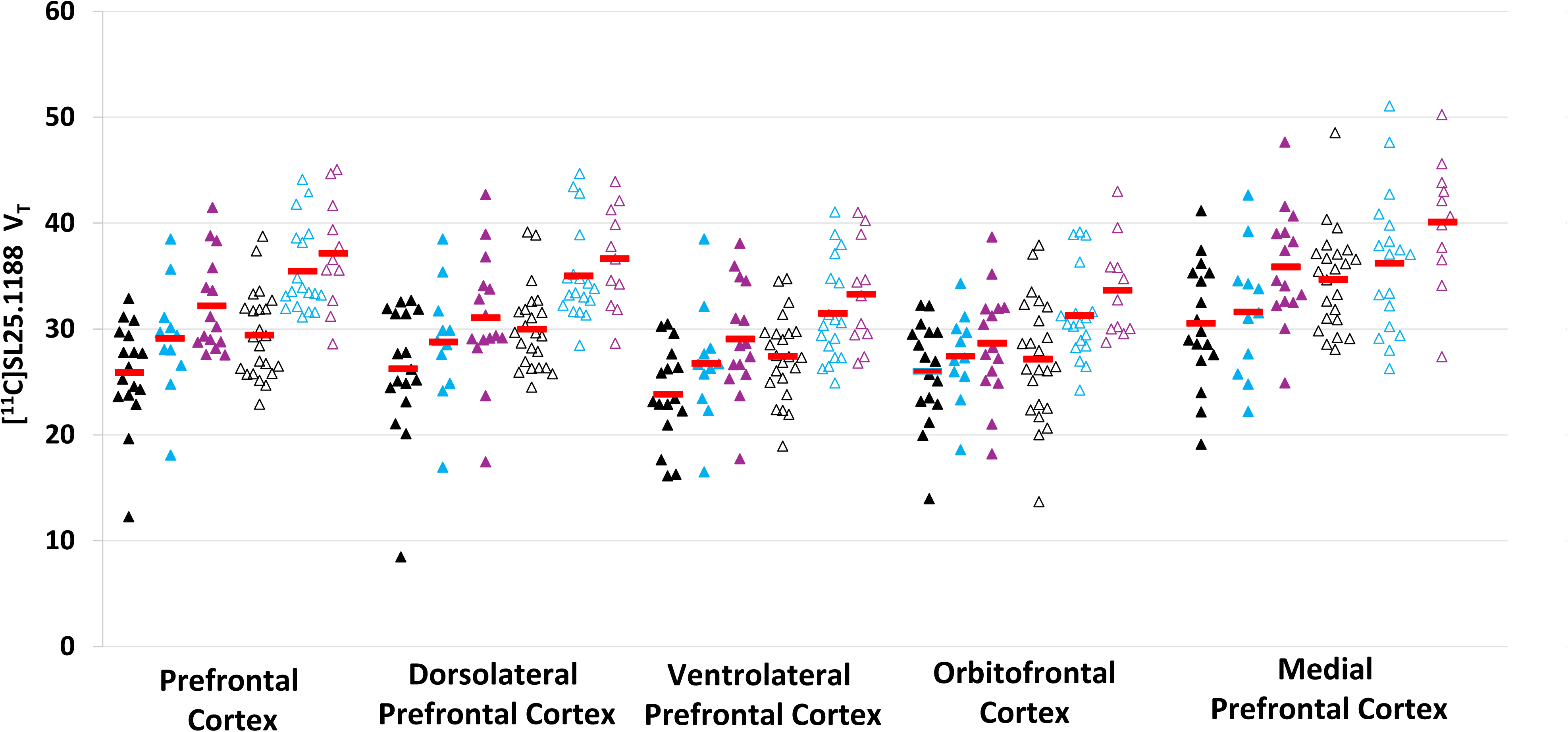
[^11^C]SL25.1188 total distribution volume (V_T_) in brain regions of interest in participants with NTR-MDE vs. TRD vs. healthy controls. A linear mixed-effects model found an effect of group on [^11^C]SL25.1188 V_T_, with greater [^11^C]SL25.1188 V_T_ in PFC subregions in TRD compared to NTR-MDE (*F*_1,52_ = 4.45, *p* = .040) and healthy controls (*F*_1,63_ = 19.23*, p* < 0.001). Black triangles represent healthy controls (*n* = 40). Blue triangles represent NTR-MDE participants (*n* = 29). Purple triangles represent TRD (*n* = 26). Filled triangles represent males (*n* = 43). Hollow triangles represent females (*n* = 52). Red horizontal bars represent means.

### Change in [^11^C]SL25.1188 V_T_ after treatment

Compliance was high, with 100%, 97.5%, and 99.6% of the medication taken on pill counts for rasagiline, phenelzine and duloxetine respectively.

When including all participants in the LMM, including treatment, region, and medication group, there was an effect of medication by treatment (*F*_2,57_ = 84.63, *p* <.001). When analyzing each individual treatment, [^11^C]SL25.1188 V_T_ was significantly lower in the PFC, ACC, dorsal putamen, ventral striatum, thalamus and hippocampus after treatment with rasagiline or phenelzine (LMM; rasagiline: *F*_1,10_ = 502.547, *p* < 0.001; phenelzine: *F*_1,7_ = 1091.052, *p* < 0.001; Fig. 3 and Table S3). The residuals in the model for the data of each treatment were not normally distributed, so results were confirmed with ANOVA within individual regions (Table S3). The LMM comparing [^11^C]SL25.1188 V_T_ before and after participants took duloxetine found an effect of treatment (*F*_1,18_ = 8.439, *p* =.010; Fig. 3), but none of the ANOVAs within individual regions had uncorrected p values below 0.05, reflecting that the change after duloxetine was modest. Mean occupancy values, calculated across brain regions for each participant using a Lassen plot, was 95.4% for rasagiline (95% CI (92.15, 98.60)), and 93.7% for phenelzine (95% CI (86.19, 101.28)).

**Figure 3:**
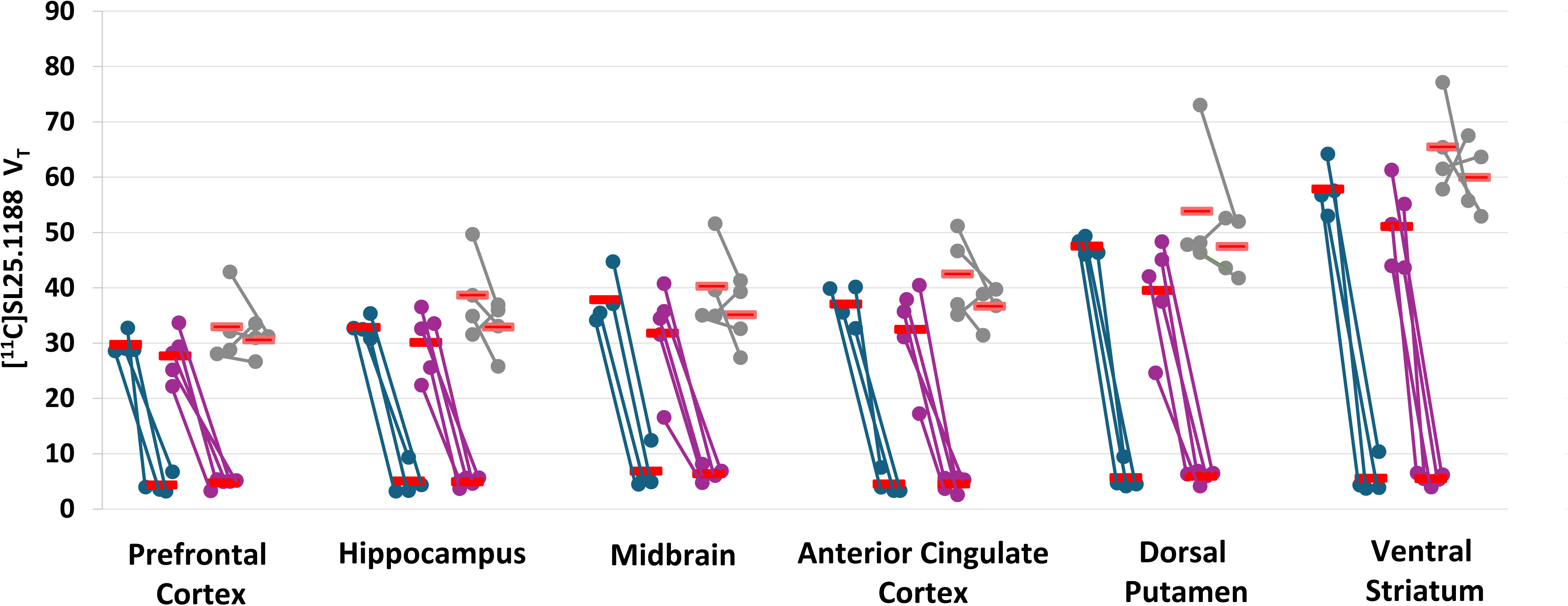
[^11^C]SL25.1188 total distribution volume (V_T_) in brain regions of interest in participants with MDE pre- and post-treatment. A linear mixed-effects model including treatment, region, and medication group found an effect of medication by treatment (*F*_2,57_ = 84.630, *p* <.001), reflecting greater effect of phenelzine and rasagiline compared to duloxetine. Blue circles represent participants who took phenelzine (*n* = 4). Purple circles represent participants who took rasagiline (*n* = 5). Grey circles represent participants who took duloxetine (*n* = 4). Red horizontal lines represent group means.

### Consistency of imaging agent administered, and free fraction across groups analyzed

[^11^C]SL25.1188 was of high radiochemical purity, with a minimum threshold of 95% and high specific activity (mean ± SD = 76.40 ± 42.18 TBq/mmol at the time of injection). There was no significant difference in specific activity between the two sexes (ANOVA, F_1,93_ = .838, p = .362) or diagnosis (ANOVA, F_2,92_ = 1.10, p = .338). There was also no significant difference between % free fraction between the two sexes (ANOVA, F_1,86_ = .951, p = .332) or across diagnostic subgroups (ANOVA, F_2,84_ = .003, p = .997) (Table S4).

### Exploration of Sex Differences in mRNA

None of the exploratory comparisons of mRNA across sex in the previously published microarray dataset yielded results at an uncorrected significance of less than 0.05 (Table S5).

## Discussion

To our knowledge, this is the first study to report sex differences in MAO-B levels in human brain. Arguably, the most novel finding was that females had significantly higher [^11^C]SL25.1188 V_T_ compared to males, which influences our understanding of sex differences for neuropsychiatric illnesses in which differential MAO-B level is implicated in modulating their pathophysiology. To our awareness, this is also the first study to investigate MAO-B in TRD, and we found significantly greater [^11^C]SL25.1188 V_T_ in the PFC in TRD compared to NTR-MDE and healthy controls, prompting consideration for targeting MAO-B in treatment resistance. Finally, we also found that rasagiline and phenelzine, in contrast to duloxetine, lowered [^11^C]SL25.1188 V_T_ across grey matter regions; and that rasagiline and phenelzine have high MAO-B occupancy. This confirms high potency of phenelzine and rasagiline at the treatment doses in clinical settings, pointing to the opportunity for future clinical development of rasagiline.

Our finding of greater [^11^C]SL25.1188 V_T_ in females was unexpected and has potential implications for sex-related differences in neuropsychiatric illnesses. MAO-B has a critical role in promoting GFAP-labelled astrogliosis, since overexpression of MAO-B promotes astrogliosis, knockdown or blockade of MAO-B reduces astrogliosis and MAO-B expression is often increased during astrogliosis [12–15]. A general, albeit simplified literature perspective is that acute astrogliosis tends to have protective advantages whereas chronic astrogliosis tends to be harmful [40, 41]. Illnesses such as Alzheimer’s disease and long COVID have higher prevalence in females [42, 43], and chronic traumatic brain injury symptoms have a longer time course in females [44]. Chronic astrogliosis with heightened level of MAO-B has been raised as an aggravating factor in the pathophysiology of all of these illnesses. We also note that greater MAO-B level can occur independently of astrogliosis, and that greater MAO-B binding was identified in the PFC of MDD, another illness that has a higher prevalence ratio of females to males. In regards to the mechanism for sex-related differences, the present finding is opposite to the original hypothesis that differential estrogen level between sex would account for a lower MAO-B level in females. Another possibility for further study is that given the MAO-B gene is located on the X chromosome, it is possible that it escapes X-chromosome inactivation. A previous report indicated that females have both greater MAO-A and MAO-B mRNA expression in grey matter throughout their lifespan [45]. However, it is acknowledged that greater mRNA level does not necessarily translate to measures of greater protein level [45], as we previously found that [^11^C]harmine V_T_, an index of MAO-A level, did not differ between males and females prior to age 41 outside of the immediate postpartum period [46–48]. Also, an exploratory analysis of a publicly available microarray dataset derived from postmortem human brain tissue yielded no difference in mRNA for MAO-B, or mRNA corresponding to markers of astrogliosis or mitochondrial density within the hippocampus, dorsolateral prefrontal cortex, or associative striatum. This raises the possibility of sex differences in MAO-B translation or stability as other explanations for sex related differences (see Supplementary Discussion for further detail).

Our finding that individuals with TRD have elevated PFC [^11^C]SL25.1188 V_T_ compared to NTR-MDE as well as healthy controls raises the question as to whether greater PFC MAO-B level may contribute to treatment resistance. Elevated MAO-B level beyond healthy control level is implicated in causing MDE symptoms, as generalized overexpression of MAO-B in mice leads to reduced locomotor speed and activity, shorter distances traveled and decreased time spent in motion in open field assessments [12]. Also, as presented in the introduction, sequelae of greater MAO-B level led to greater removal of non-serotoninergic monoamines, greater production of reactive oxygen species and loss of mitochondrial complex I [11, 12], processes also implicated in causing depressive symptoms. The present study also demonstrated that [^11^C]SL25.1188 V_T_ is not substantively affected by SNRI treatment. So, given that MAO-B level is not well targeted by SNRIs, is more prominently elevated in TRD, and elevation in MAO-B level from the usual state predisposes towards symptoms and sequelae that may contribute to depressive symptoms, it may be worth assessing whether MAO-B inhibitors may have therapeutic efficacy in TRD.

Non-selective MAO inhibitors like phenelzine have some evidence for benefit in clinical trials for treatment resistant depression [32], but as the rationale was largely ascribed to MAO-A inhibition [11], selective MAO-B inhibitors have not had similar investigation. Non-selective MAO inhibitor tranylcypromine was assessed in treatment resistance in a STAR*D trial, but unfortunately, poor tolerability of tranylcypromine in that trial led to high non-completion rates and dosing that may have been lower than optimal for treatment response [34, 49]. It is also uncertain whether the relatively low dose used in that trial achieved a substantive MAO-B occupancy [34].

Both phenelzine and rasagiline caused high magnitude reductions of [^11^C]SL25.1188 V_T_ with occupancy of 94 to 95 per cent in contrast to duloxetine. One implication is that phenelzine at a total daily dose of 45 to 60mg is associated with substantive occupancy, suggesting that such dose is often adequate for target engagement of MAO-B inhibition. A second implication is that high occupancy of rasagiline, being similar to the antidepressant phenelzine, argues for consideration of development of rasagiline to target MAO-B in mood disorders, moving beyond its approvals presently obtained only for Parkinson’s disease. For example, there are reports that rasagiline improves depression outcomes in Parkinson’s disease even when controlling for motor function [50, 51], and decreases avolition, a symptom also commonly found in MDD [52]. A third consideration is that new and more selective MAO-B inhibitors in development [4, 5] should consider the high occupancies of phenelzine and rasagiline as comparative, achievable thresholds when determining occupancy for therapeutic trials.

There are some limitations to the present study, some common to PET imaging investigations. First, [^11^C]SL25.1188 V_T_ reflects both specific binding of the radiotracer to MAO-B and non-specific binding. However, based on the Lassen plots, 88 to 95 per cent of V_T_ is represented by specific binding, so changes in non-displaceable binding are unable to account for sex differences or effects of phenelzine or rasagiline, and would require large improbable changes to account for the effect of treatment resistance. Second, while typical of PET occupancy studies, the number of participants enrolled in the treatment component of the study was low in the study plan because the variability in the blocked condition is small. Third, the magnitude of difference in [^11^C]SL25.1188 V_T_, while substantive comparing TRD to healthy, was modest in the comparison of TRD to NTR-MDE, so the notion of investigating MAO-B inhibitors as clinical interventions in TRD presented earlier might only be relevant to a subset of TRD, requiring a stratification approach in future investigations.

In conclusion, the main findings were greater [^11^C]SL25.1188 V_T_ in females and in TRD, and that both phenelzine and rasagiline strongly reduced [^11^C]SL25.1188 V_T_, unlike duloxetine. The interpretation of greater MAO-B level in brain grey matter in females raises important implications as to whether this contributes to sex differences observed in illnesses with prominent MAO-B-labelled astrogliosis like Alzheimer’s disease, traumatic brain injury or long COVID and/or illnesses with only elevated MAO-B level like major depressive disorder. The interpretation that greater MAO-B level occurs in TRD, combined with lack of effect of SNRI duloxetine and roles of greater MAO-B level in predisposition to symptoms of depression, raises consideration for greater development of well-tolerated MAO-B inhibitors with substantial occupancy for TRD. Consistent with this direction, we demonstrate that the occupancy for two MAO-B inhibitor medications with clinical indications is high, and comparable across less selective MAO-B inhibitor phenelzine and better tolerated, more selective MAO-B inhibitor rasagiline.

## Data Availability Statement

The data for the main figures and graphs of this study are available within the supplementary material.

## Acknowledgments

The authors acknowledge the valuable contributions of the study participants as well as research staff at the Centre for Addiction and Mental Health.

## Author Contributions

Dr. Jeffrey Meyer and Emily Kuik had full access to all of the data in the study and take responsibility for the integrity of the data and the accuracy of the data analysis.

*Concept and design:* Meyer, Rusjan, Bagby.

*Acquisition, analysis, or interpretation of data:* Kuik, Miler, Baena-Tan, Moriguchi, Rusjan, Husain, Kloiber, Braga, Meyer.

*Drafting of the manuscript:* Kuik, Meyer.

*Critical revision of the manuscript for important intellectual content:* Kuik, Baena-Tan, Moriguchi, Braga, Miler, Rusjan, Bagby, Husain, Vasdev, Tomoda, Banasr, Kloiber, Meyer

*Statistical analysis:* Kuik, Miler, Baena-Tan, Meyer.

*Obtained funding:* Rusjan, Bagby, Vasdev, Meyer.

*Administrative, technical, or material support:* Rusjan.

*Supervision:* Meyer.

