## Supplementary Materials for "Effects of sex, treatment resistance, and antidepressant treatment on monoamine oxidase B total distribution volume"

Kuik *et al.*

Table of Contents

**Supplemental Methods**	1

**Supplemental Discussion**	6

**Table S1.** Regional [^11^C]SL25.1188 V_T_ in TRD and NTR-MDE	8

**Table S2.** Regional [^11^C]SL25.1188 V_T_ in TRD and	controls	9

**Table S3.** Regional [^11^C]SL25.1188 V_T_ pre- and post-treatment	10

**Table S4.** [^11^C]SL25.1188 PET scan parameters of main participant groups	11

**Table S5.** Comparing mRNA levels between females and males	12

### Supplemental Methods

#### Participants

Participants were scanned between September 2012 and April 2024. All participants met additional exclusion criteria required for PET scanning, including not pregnant or breastfeeding; no blood coagulation disorders or ongoing use of anticoagulant medication, no claustrophobia; weight and height not exceeding the capacity of PET and MRI scanners (400lbs and 7ft); and not exceeded a total annual radiation dose of 20mSv or have completed 8 or more PET scans during their lifetimes. Participants are also required to not have any metal objects or implanted electrical devices in the body that would preclude MRI scanning.

Participants who completed the open trial treatment portion of the study also met additional exclusion criteria: not positive for hepatic dysfunction, either by history or measured by aspartate aminotransferase (AST) and alanine aminotransferase (ALT) tests; no diagnosis of cardiovascular disease such as hypertension/hypotension, angina or tachycardia. Participants in the duloxetine group also had an exclusion criterion of no history of non-response to duloxetine at a daily dose of 60mg daily or higher.

Participants in either the phenelzine or rasagiline groups met additional inclusion criteria, including history of non-response to serotonin reuptake inhibitor medication and non-response to a medication that raises extracellular norepinephrine; or history of non-response to a medication that raises extracellular serotonin and norepinephrine. Participants were also required to have either a history of non-response to bupropion or lithium, or did not wish to take these treatments. Participants were also aware and willing to follow medication and/or dietary requirements required for taking phenelzine or rasagiline depending on their treatment group.

Participants in either the phenelzine or rasagiline group also met additional exclusion criteria, including no previous hypersensitivity to monoamine oxidase inhibitors or tyramine; no previous diagnosis of cerebrovascular or cardiovascular disorders, or recurrent or frequent headaches; no diagnosis of phaeochromocytoma and catecholamine-releasing paragangliomas; and no use of triptans or tryptamines (e.g., sumatriptan or rizatriptan) in the past 2 weeks. Systolic blood pressure was not outside of 91 to 139 mmHg (inclusive), diastolic blood pressure was not outside of 51 to 90 mmHg (inclusive), and there was no decrease in systolic blood pressure of 20 mm Hg or diastolic blood pressure of 10 mm Hg within three minutes of standing when compared with BP from the sitting position.

#### PET and MRI Scanning

A transmission scan measured using a single photon point source, 137Cs (t_1/2_=30.2 years, Eγ=662 keV) was acquired immediately before the acquisition of the emission scan. For the emission scan, [^11^C]SL25.1188 was infused intravenously over a 30-second period at a constant rate using a Harvard infusion pump (Harvard Apparatus). Scanning time was 90 minutes with the following frame times: 5 × 30 seconds, 1 × 45 seconds, 2 × 60 seconds, 1 × 90 seconds, 1 × 120 seconds, 1 × 210 seconds, and 16 × 300 seconds [1]. Attenuation correction was completed with a 137Cs transmission scan acquired in 64 bit list mode, which was converted into a 511 keV attenuation correction image [2]. Emission images were acquired in 64 bit list mode, and were later reconstructed from 3D sinograms. Key steps in reconstruction include accounting for the octagonal design of the tomograph [3], correction for photon attenuation, detector normalization, and scatter in the 3D sinogram [4]; fourier rebinning to convert 3D into 2D sinograms [5]; 3 reconstruction into image space using a 2D filtered back projection algorithm with a HANN filter at Nyquist cut-off frequency and calibration of the images to nCi/cc. Using the automatic blood sampling system (ABSS, Model # PBS-101 from Veenstra Instruments, Joure, Netherlands), arterial sampling was taken continuously for the first 22 minutes starting at a rate of 350 ml/hr, then slowed to 150 ml/hr after 7.25 min. Manual samples of 5 to 10 ml were taken before and during scanning, at t= 3.5, 7, 12, 15, 20, 30, 45, 60, and 80 minutes at volumes of 5, 5, 5, 5, 10, 10, 10, 10, and 10 ml. The delay, dispersion and metabolite corrected input function were created as previously described [1].

Magnetic resonance imaging (MRI) scans were acquired on a GE Discovery MR750 3T scanner (General Electric Medical Systems, Milwaukee, WI) equipped with an 8-channel headcoil at the Brain Health Imaging Centre at CAMH. T1-weighted MRI acquired in 3D of slice thickness 0.9mm were obtained (TR 6.7ms, TE 3.0ms, flip angle 8°, NEX=1, acquisition matrix 256 x 256; FOV 23cm, duration 4 min 42 s). Regions of Interest were delineated using in-house image analysis software ROMI [6]. In brief, a standard brain template (International Consortium for Brain Mapping/Montreal Neurological Institute 152 MRI) containing a set of predefined cortical and subcortical ROIs (based upon the neuroanatomy atlas of Duvernoy [7] but consistent with the atlas of Talairach [8]; the division of the striatum is from Mawlawi et al. [9] and the prefrontal cortex subregions are derived from the cytoarchitectural definitions of Rajkowska) [10-12] was used. These ROI are transformed and deformed (SPM normalization; Wellcome Dept. of Cognitive Neurology, London, UK; http://www.fil.ion.ucl.ac.uk/spm/) to fit each individual high-resolution proton density-weighted MRI. Each individual's set of automatically created ROIs is then refined by iteratively including and deleting voxels based on the probability of each voxel belonging to grey matter (SPM8 segmentation, Wellcome Department of Cognitive Neurology, London, UK; http://www.fil.ion.ucl.ac.uk/spm). Each MRI is co-registered to the summed PET image using normalized mutual information algorithm implemented under SPM8. The resulting transformation is applied to the individual's refined ROIs and then resliced to match the dimensions of the PET images. The location of the ROI is verified by visual assessment of the ROI on the co-registered MRI and summated [^11^C]SL25.1188 PET image. Time activity curves are visually inspected and, if evidence for motion artifact is present, a motion correction algorithm is applied. Head movement in the dynamic PET acquisition is corrected using frame-by-frame realignment. A normalized mutual information algorithm is applied with SPM8 (Wellcome Trust Centre for Neuroimaging, London, UK) to co-register each frame to the 12th frame after the radiopharmaceutical arrives to the field of view, which shows a high signal-to-noise ratio. Parameters from the normalized mutual information are then applied to the corresponding attenuation-corrected dynamic images to generate a movement-corrected dynamic image. The two tissue compartment analysis, previously validated for this radiotracer [1], was performed using PMOD Kinetic Modeling Tool (PKIN) version 4·2 (PMOD technologies, Zurich, Switzerland) to measure [^11^C]SL2511.88 total distribution volume ([^11^C]SL2511.88 V_T_). All statistical analyses were conducted using IBM SPSS Statistics for Windows version 25·0 (IBM Corp., Armonk, N.Y., USA).

#### Exploration of Sex Differences in mRNA

Microarray data obtained from [GSE53987](https://ncbi.nlm.nih.gov/geo/query/acc.cgi?acc=GSE53987) was used to explore mechanisms involved in the observed sex differences in our study. This dataset contained samples from the hippocampus, dorsolateral prefrontal cortex, and associative subregion of dorsal striatum. After verifying that samples between females and males did not differ significantly by age, pH, RNA Integrity Number (RIN), or post-mortem interval (PMI), differential gene expression between males and females was performed using the GEO2R web tool, which implements the Bioconductor *limma* package [13].

Gene expression of MAO-B, EAPP and R1 were explored to assess whether potential sex differences in [^11^C]SL2511.88 V_T_ might be attributable to differences in the transcription of MAO-B or the transcription regulators. In addition, as GFAP and citrate synthase levels are known to vary with astrogliosis and mitochondrial density respectively, sex difference in the relationship between these genes and MAO-B levels were also explored.

For the mRNA comparisons between sexes, a moderated t-test was used to compare the difference between female and male samples. P values were adjusted using the Benjamini–Hochberg false discovery rate (FDR) procedure to correct for multiple comparisons.

### Power

The effect size of history of treatment resistance versus not having a history of treatment resistance on PFC [^11^C]SL25.1188 V_T_ was anticipated to be 0.8 to 1. With an α coefficient of 0.05, the power is greater than 80% when comparing samples of 26 versus 26.

The mean difference in test-retest data measurement of [^11^C]SL25.1188 V_T_ is zero with a standard deviation of approximately 2.5 [^11^C]SL25.1188 V_T_. For an expected reduction of 12 [^11^C]SL25.1188 V_T_ or greater, corresponding to an occupancy of 65%, power is greater than 95% to detect a change in a group of approximately 6 participants.

### Supplemental Discussion

While we have raised escape of X chromosome inactivation as a possible mechanism to account for differential [^11^C]SL25.1188 V_T_, the absence of sex differences in mRNA of MAO-B suggests a contrary position that the observed sex difference in [^11^C]SL25.1188 V_T_ is not driven by differential level of MAO-B mRNA. Given this lack of sex difference in MAO-B mRNA, it could be consistent that there is also no sex difference in mRNA of EAPP (E2F-associated phosphoprotein) and R1, the latter two being proteins which regulate MAO-B transcription. To our knowledge, sex specific differences in glial fibrillary acidic protein (GFAP) labelled astrogliosis (which may have elevated expression of MAO-B protein) have not been reported. A difference in mRNA levels of GFAP would have suggested consideration of differential GFAP-labelled astrogliosis to account for sex-related differences in this sample, which was not found. Citrate synthase activity is sometimes applied as a marker of mitochondrial density and citrate synthase mRNA did not differ by sex. Hence, this line of investigation did not suggest future investigation of differences in mitochondrial density across sex as an explanation for differential [^11^C]SL25.1188 V_T_. Lack of sex difference in MAO-B mRNA may seem inconsistent with MAO-B protein differences. However, discrepancies between mRNA and protein levels can occur as a result of processes such as autophagy as well as alterations in transcription or translation in response to greater oxidation and/or stress [14, 15]. While none of our explorations evaluated protein level (or citrate synthase activity), they may suggest that future study of sex related differences in [^11^C]SL25.1188 V_T_ should consider differential translation and/or stability of MAO-B protein.

**Table S1.** Regional [^11^C]SL25.1188 V_T_ in TRD and NTR-MDE.

|  | **TRD** (*n* = 26) | | **NTR-MDE**  (*n* = 29) | |  |  |  |
| --- | --- | --- | --- | --- | --- | --- | --- |
| **Region of Interest** | **Mean** | **SD** | **Mean** | **SD** | **Difference** | ***F*** | ***p*** |
| Prefrontal Cortex | 34.3 | 5.4 | 32.3 | 5.6 | 6.0% | 4.778 | .033 |
| Dorsolateral Prefrontal Cortex | 33.4 | 6.2 | 31.9 | 5.9 | 4.6% | 2.963 | .091 |
| Ventrolateral Prefrontal Cortex | 30.8 | 5.5 | 29.1 | 5.5 | 5.7% | 3.236 | .078 |
| Medial Prefrontal Cortex | 37.7 | 6.0 | 34.5 | 6.8 | 8.9% | 5.807 | .020 |
| Orbitofrontal Cortex | 30.8 | 5.5 | 29.2 | 4.6 | 5.3% | 3.348 | .073 |
| Anterior Cingulate Cortex | 40.8 | 7.1 | 38.1 | 6.6 | 6.8% | 4.527 | .038 |
| Hippocampus | 37.3 | 6.3 | 36.3 | 5.1 | 2.7% | 1.623 | .208 |
| Dorsal Putamen | 50.5 | 7.6 | 49.5 | 7.8 | 2.0% | 1.122 | .294 |
| Ventral Striatum | 60.4 | 11.2 | 58.3 | 8.9 | 3.5% | 1.513 | .224 |
| Thalamus | 57.2 | 8.9 | 54.8 | 9.4 | 4.2% | 1.913 | .173 |
| Midbrain | 38.1 | 5.2 | 37.6 | 6.8 | 1.3% | 0.449 | .506 |
| Parietal Cortex | 33.0 | 4.9 | 31.3 | 4.1 | 5.3% | 3.699 | .060 |
| Temporal Cortex | 33.2 | 4.1 | 31.6 | 5.1 | 4.9% | 2.962 | .091 |
| Occipital Cortex | 28.4 | 3.7 | 26.8 | 4.9 | 5.8% | 4.743 | .034 |
| Means and standard deviations presented in table are derived from raw unadjusted V_T_ values. P values are derived from an analysis of covariance (ANCOVA) with sex as covariate. | | | | | | | |

**Table S2.** Regional [^11^C]SL25.1188 V_T_ in TRD and controls.

|  | **TRD** (*n* = 26) | | **Controls**  (*n* = 40) | |  |  |  |
| --- | --- | --- | --- | --- | --- | --- | --- |
| **Region of Interest** | **Mean** | **SD** | **Mean** | **SD** | **Difference** | ***F*** | ***p*** |
| Prefrontal Cortex | 34.3 | 5.4 | 27.9 | 4.8 | 22.9% | 35.388 | <.001 |
| Dorsolateral Prefrontal Cortex | 33.4 | 6.2 | 28.4 | 5.2 | 17.6% | 18.588 | <.001 |
| Ventrolateral Prefrontal Cortex | 30.8 | 5.5 | 25.9 | 4.5 | 18.9% | 22.468 | <.001 |
| Medial Prefrontal Cortex | 37.7 | 6.0 | 32.9 | 5.5 | 14.6% | 15.283 | <.001 |
| Orbitofrontal Cortex | 30.8 | 5.5 | 26.7 | 5.4 | 15.4% | 11.173 | .001 |
| Anterior Cingulate Cortex | 40.8 | 7.1 | 35.5 | 5.8 | 14.9% | 15.578 | <.001 |
| Hippocampus | 37.3 | 6.3 | 32.5 | 6.8 | 14.8% | 13.996 | <.001 |
| Dorsal Putamen | 50.5 | 7.6 | 44.8 | 7.9 | 12.7% | 13.946 | <.001 |
| Ventral Striatum | 60.4 | 11.2 | 54.6 | 14.9 | 10.6% | 4.456 | .039 |
| Thalamus | 57.2 | 8.9 | 48.5 | 9.0 | 17.9% | 20.184 | <.001 |
| Midbrain | 38.1 | 5.2 | 34.5 | 8.0 | 10.4% | 5.840 | .019 |
| Parietal Cortex | 33.0 | 4.9 | 28.6 | 4.5 | 15.4% | 17.707 | <.001 |
| Temporal Cortex | 33.2 | 4.1 | 29.2 | 5.8 | 13.7% | 11.055 | .001 |
| Occipital Cortex | 28.4 | 3.7 | 24.5 | 4.0 | 15.9% | 21.003 | <.001 |
| Means and standard deviations presented in table are derived from raw unadjusted V_T_ values. P values are derived from an analysis of covariance (ANCOVA) with sex as covariate. | | | | | | | |

**Table S3.** Regional [^11^C]SL25.1188 V_T_ pre- and post-treatment.

| **Region of Interest** | **Treatment** | **Pre-treatment** | | **Post-treatment** | | **Group Difference** | | |
| --- | --- | --- | --- | --- | --- | --- | --- | --- |
|  |  | **Mean** | **SD** | **Mean** | **SD** | **Difference** | ***F*** | ***p*** |
| Prefrontal Cortex | Phenelzine | 29.8 | 2.0 | 4.4 | 1.6 | 85.4% | 400.86 | <.001 |
|  | Rasagiline | 26.9 | 5.8 | 4.7 | 0.8 | 82.4% | 72.02 | <.001 |
|  | Duloxetine | 32.9 | 6.9 | 30.6 | 2.9 | 7.2% | 0.41 | 0.547 |
| Medial Prefrontal Cortex | Phenelzine | 34.3 | 1.7 | 5.1 | 1.9 | 85.2% | 545.07 | <.001 |
|  | Rasagiline | 31.5 | 6.0 | 4.8 | 0.8 | 84.9% | 97.49 | <.001 |
|  | Duloxetine | 38.2 | 6.7 | 34.9 | 3.0 | 8.6% | 0.79 | 0.407 |
| Dorsolateral Prefrontal Cortex | Phenelzine | 29.5 | 1.8 | 4.5 | 1.6 | 84.6% | 415.87 | <.001 |
|  | Rasagiline | 26.8 | 6.0 | 4.6 | 0.8 | 82.9% | 66.85 | <.001 |
|  | Duloxetine | 32.6 | 7.0 | 31.0 | 2.6 | 4.9% | 0.19 | 0.682 |
| Ventrolateral Prefrontal Cortex | Phenelzine | 27.9 | 1.4 | 4.5 | 1.8 | 83.8% | 429.82 | <.001 |
|  | Rasagiline | 23.6 | 4.5 | 5.1 | 1.4 | 78.2% | 78.36 | <.001 |
|  | Duloxetine | 29.1 | 6.9 | 27.3 | 3.9 | 6.4% | 0.22 | 0.653 |
| Orbitofrontal Cortex | Phenelzine | 28.1 | 2.4 | 4.2 | 1.8 | 85.1% | 255.35 | <.001 |
|  | Rasagiline | 25.6 | 4.8 | 4.5 | 0.9 | 82.6% | 95.36 | <.001 |
|  | Duloxetine | 30.6 | 6.1 | 27.4 | 3.0 | 10.5% | 0.90 | 0.380 |
| Anterior Cingulate Cortex | Phenelzine | 37.1 | 3.6 | 4.5 | 2.0 | 87.8% | 248.58 | <.001 |
|  | Rasagiline | 33.4 | 7.3 | 4.6 | 1.4 | 86.4% | 75.04 | <.001 |
|  | Duloxetine | 42.5 | 7.7 | 36.7 | 3.7 | 13.7% | 1.85 | 0.223 |
| Hippocampus | Phenelzine | 32.8 | 1.9 | 5.1 | 2.9 | 84.6% | 258.55 | <.001 |
|  | Rasagiline | 31.5 | 4.2 | 4.9 | 0.8 | 84.4% | 191.76 | <.001 |
|  | Duloxetine | 38.7 | 7.9 | 32.9 | 5.0 | 14.9% | 1.52 | 0.264 |
| Dorsal Putamen | Phenelzine | 47.5 | 1.6 | 5.7 | 2.5 | 88.0% | 796.72 | <.001 |
|  | Rasagiline | 41.6 | 5.4 | 6.0 | 1.1 | 85.7% | 213.34 | <.001 |
|  | Duloxetine | 53.8 | 12.8 | 47.5 | 5.6 | 11.8% | 0.83 | 0.398 |
| Ventral Striatum | Phenelzine | 57.9 | 4.6 | 5.6 | 3.2 | 90.4% | 342.93 | <.001 |
|  | Rasagiline | 51.3 | 7.3 | 5.5 | 1.0 | 89.3% | 194.29 | <.001 |
|  | Duloxetine | 65.5 | 8.4 | 60.0 | 6.8 | 8.4% | 1.05 | 0.346 |
| Thalamus | Phenelzine | 55.9 | 1.9 | 6.5 | 3.7 | 88.4% | 556.61 | <.001 |
|  | Rasagiline | 47.3 | 9.2 | 6.3 | 1.2 | 86.7% | 96.72 | <.001 |
|  | Duloxetine | 57.8 | 12.9 | 49.9 | 4.2 | 13.6% | 1.34 | 0.291 |
| Midbrain | Phenelzine | 37.9 | 4.7 | 6.9 | 3.7 | 81.8% | 105.88 | <.001 |
|  | Rasagiline | 34.6 | 4.0 | 6.4 | 1.2 | 81.5% | 224.38 | <.001 |
|  | Duloxetine | 40.3 | 7.9 | 35.1 | 6.4 | 12.8% | 1.04 | 0.347 |
| Parietal Cortex | Phenelzine | 30.3 | 2.6 | 5.0 | 1.9 | 83.7% | 248.69 | <.001 |
|  | Rasagiline | 28.0 | 5.8 | 5.5 | 1.6 | 80.2% | 69.8 | <.001 |
|  | Duloxetine | 32.8 | 8.1 | 30.1 | 3.1 | 8.2% | 0.38 | 0.559 |
| Temporal Cortex | Phenelzine | 29.4 | 2.6 | 4.7 | 1.7 | 83.8% | 254.98 | <.001 |
|  | Rasagiline | 28.4 | 4.8 | 5.0 | 0.8 | 82.5% | 116.6 | <.001 |
|  | Duloxetine | 32.8 | 7.7 | 29.3 | 4.2 | 10.9% | 0.65 | 0.449 |
| Occipital Cortex | Phenelzine | 25.4 | 3.0 | 4.6 | 1.9 | 81.8% | 136.29 | <.001 |
|  | Rasagiline | 25.3 | 5.4 | 5.0 | 1.1 | 80.3% | 69.06 | <.001 |
|  | Duloxetine | 28.9 | 7.7 | 26 | 4.8 | 10.2% | 0.420 | 0.540 |

Means and standard deviations presented in table are derived from raw unadjusted V_T_ values.

P values are derived from an analysis of variance (ANOVA).

**Table S4.** [^11^C]SL25.1188 PET scan parameters of main participant groups

|  | **Injected Activity (MBq)** | | **Specific Activity (TBq/mmol)** | | **Free Fraction (%)** | |
| --- | --- | --- | --- | --- | --- | --- |
| **Group** | **Mean** | **SD** | **Mean** | **SD** | **Mean** | **SD** |
| TRD (n=26) | 352.4 | 26.9 | 80.8 | 38.4 | 1.7 | 0.4 |
| NTR-MDE (n=29) | 350.2 | 36.9 | 76.0 | 37.3 | 1.8^i^ | 0.4^i^ |
| Controls (n=40) | 358.4 | 24.9 | 69.8 | 40.6 | 1.8 | 0.4 |

ANOVA did not find a significant difference in specific activity (F_2,92_ = 1.10, p = .338) or % free fraction (*F*_2,84_ = .003, *p* = .997) across diagnostic subgroups.

^i^Two NTR-MDE blood samples were not available for free fraction analysis.

**Table S5.** Comparing mRNA levels between females and males

| **Gene name** | **t** | **B** | **Log2-fold change** | **p value** | **adjusted p value** |
| --- | --- | --- | --- | --- | --- |
| Monoamine oxidase B (MAO-B) | -0.824 | -5.068 | -0.080 | 0.412 | 0.827 |
| Cell division cycle associated 7 like (CDCA7L)^i^ | -0.843 | -5.056 | -0.202 | 0.401 | 0.827 |
| E2F associated phosphoprotein (EAPP) | -0.731 | -5.124 | -0.092 | 0.467 | 0.827 |
| Glial fibrillary acidic protein (GFAP) | -0.902 | -5.016 | -0.184 | 0.369 | 0.827 |
| Citrate synthase | -0.824 | -5.068 | -0.100 | 0.412 | 0.827 |

P values are derived from a moderated t-test. Adjusted p values are adjusted for multiple testing with the Benjamini & Hochberg false discovery rate method.

^i^Also known as R1.
